# Calprotectin as a sepsis diagnostic marker in critical care: a retrospective observational study

**DOI:** 10.1101/2024.12.18.24319225

**Authors:** Maria Lengquist, Vera Sundén-Cullberg, Sofie Hyllner, Hazem Koozi, Anders Larsson, Lisa Mellhammar, Hans Friberg, Alexandru Schiopu, Attila Frigyesi

## Abstract

**Background:** Diagnosing sepsis in critical care remains a challenge due to the lack of gold-standard diagnostics. Calprotectin (S100A8/A9) has been proposed as a diagnostic marker to identify sepsis in critically ill patients. This study evaluated the diagnostic performance of calprotectin and C-reactive protein (CRP) to distinguish between sepsis and non-sepsis on intensive care unit (ICU) admission.

**Methods:** Admission biobank blood samples from adult patients admitted to four ICUs (2015-2018) were used to analyse calprotectin and CRP. All adult patients were screened retrospectively for the sepsis-3 criteria at ICU admission. The diagnostic performance of calprotectin and CRP was evaluated using receiver operating characteristic (ROC) curves.

**Results:** We included 4732 patients, of whom 44% had sepsis. Calprotectin levels were higher in sepsis (p*<*0.001). The area under the receiver operating curve (AUROC) to diagnose sepsis was 0.61 for calprotectin compared to 0.72 for CRP (p*<*0.001). Among microbiological subgroups of sepsis patients, fungal sepsis had the highest level of calprotectin.

**Conclusions:** The diagnostic performance of calprotectin in identifying sepsis patients at ICU admission was inferior to that of CRP.

## Background

Sepsis is a life-threatening condition caused by a dysregulated host response to an infection and is common among patients requiring intensive care[1, 2].

A key challenge in identifying sepsis in patients requiring intensive care is that a majority have organ failure[2]. Fulfilling the sepsis criteria in the Intensive Care Unit (*ICU*) setting is mainly dependent on infection identification.

Biomarkers can help diagnose infections and sepsis, but they need to be specific for infection and not merely reflect critical illness or systemic inflammation. While many biomarkers have been assessed for their diagnostic performance in sepsis, no single biomarker has been identified with enough sensitivity and specificity to confirm or rule out sepsis[3, 4]. Moreover, novel biomarkers should provide additional value compared to established biomarkers, such as C-reactive Protein (*CRP*), to reach clinical use[5].

Calprotectin is a complex of two proteins (S100A8 and S100A9) which can be found intracellularly in immune and endo/epithelial cells, most notably in neutrophils and monocytes[6]. Calprotectin is secreted from cells after immune activation, and rapidly increasing blood levels can be measured within hours after bacterial or endotoxin exposure[7, 8].

Previous reports indicate that calprotectin could serve as a diagnostic biomarker for bacterial sepsis[9]. The assessment of calprotectin as a marker of sepsis in the *ICU* is limited to a few small studies[10, 11]. Thus, there is a need to evaluate calprotectin in larger cohorts of critically ill patients, as well as perform comparisons with other biomarkers of inflammation, before considering further clinical use.

### Objectives

This study aimed to investigate calprotectin’s diagnostic performance in distinguishing sepsis from non-sepsis at *ICU* admission, to compare the performance of calprotectin with *CRP*, and to assess both biomarkers in subgroups of critically ill *ICU* patients.

## Methods

### Study design and setting

This is a retrospective observational sub-study of the Swecrit biobank study, conducted at four *ICU* s at Skåne University Hospital in Lund and Malmö, Helsingborg Hospital and Kristianstad Hospital between 2015-2018[12]. The Swecrit biobank study collected blood samples from adult (*≥*18 years old) patients at admission to a participating *ICU*. Blood samples were stored in the Region Scania biobank, Lund, Sweden.

The Standards for Reporting Diagnostic accuracy studies (*STARD*) guidelines were followed[13].

### Participants

This study included all adult *ICU* patients with an *ICU* Length of stay (*LOS*) of *>*24 hours. Patients deceased within 24 hours of *ICU* admission were also included. Exclusion criteria were discharge within 24 hours of *ICU* admission.

For identification of sepsis, the sepsis-3 criteria were used[1]. Our operational sepsis criteria were a Sequential Organ Failure Assessment (*SOFA*)-score*≥*2 within *±*1 hour of *ICU* admission in combination with a retrospective assessment of infection. Criteria for infection were either 1) culture positivity (see Supplement 1) within *±*48 hours of *ICU* admission, or 2) culture negativity AND suspected infection (blood culture sampling within 24 hours of *ICU* admission and administration of antibiotics[14]) AND a probable infection according to the Linder-Mellhammar Criteria of Infection (*LMCI*)[15, 16].

Culture-positive sepsis patients were assigned to one of the pre-defined microbiological subgroups (gram-positive, gram-negative, virus, fungus, polymicrobial), see Supplement 1 for definitions.

### Data sources

Clinical data, such as vital signs and variables used to calculate Simplified Acute Physiology Score 3 (*SAPS −* 3) and *SOFA* scores, were entered by the treating physician into the PASIVA software and forwarded to the Swedish Intensive Care Registry (*SIR*)[17, 18]. Laboratory values (except calprotectin and *CRP*), microbiological testing and results were automatically extracted from the hospital laboratory electronic system. Administration of antibiotics and screening of *LMCI* were based on a review of the Electronic Medical Records (*EMR*) for culture-negative cases who had blood cultures sampled. See Supplement 2 for the data sources of variables used in Table 1.

**Table 1:**
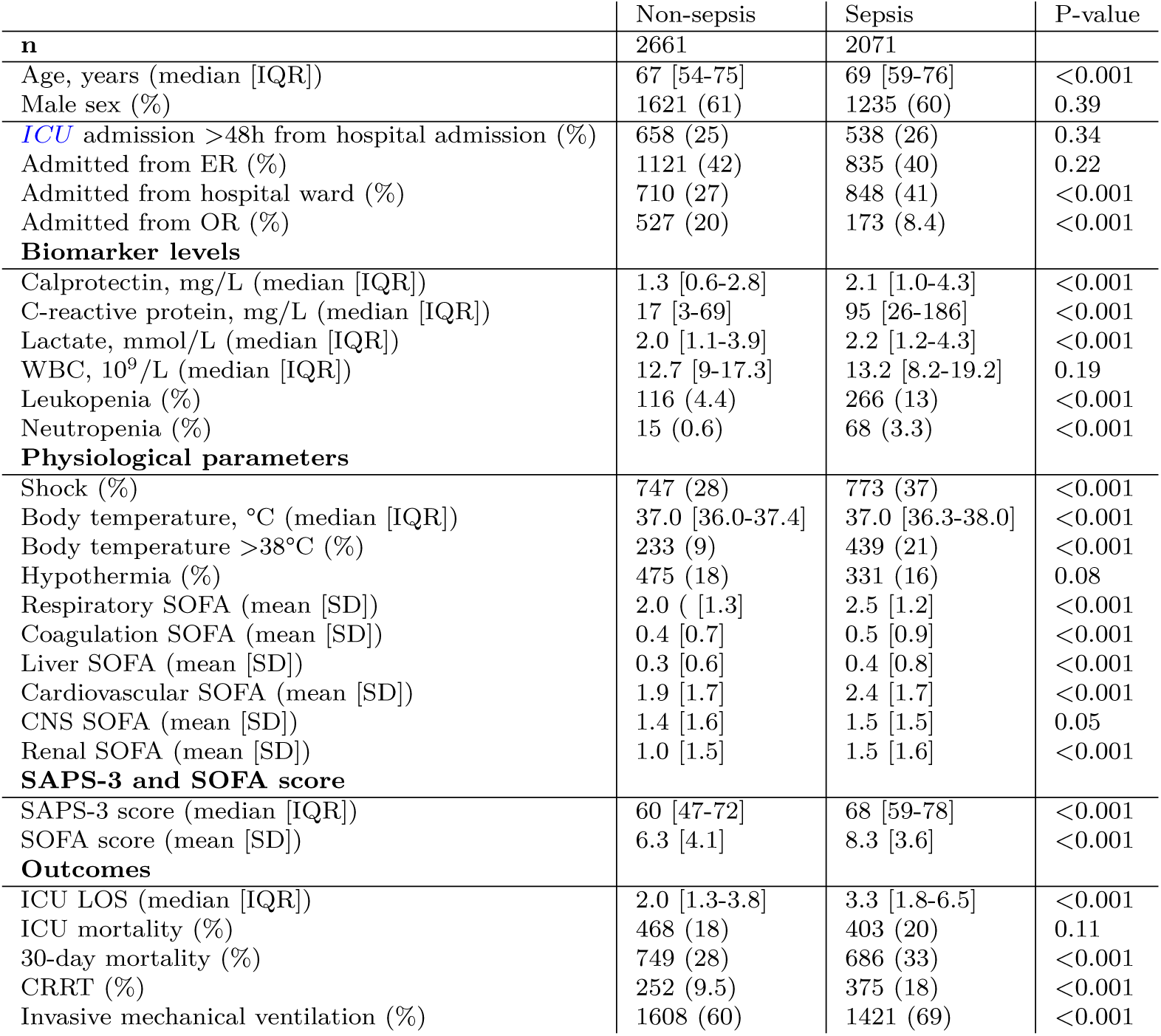
Descriptive statistics and outcomes of the non-sepsis and sepsis cohorts. All variables, except outcomes, are at *ICU* admission. *CRRT* and mechanical ventilarion are during the *ICU* stay. *ICU; intensive care unit. LOS; length of stay. ER; emergency room. OR; operating room. IQR; interquartile range. SD; standard deviation. CRP; C-reactive protein. WBC; white blood cell count. PF-ratio; PaO_2_/FiO_2_-ratio (oxygenation index). SAPS-3; simplified acute physiology score-3. SOFA; sequential organ failure assessment. CRRT; continuous renal replacement therapy*.

### Variables

For biomarkers routinely analysed at *ICU* admission, the values closest in time to *ICU* admission, but within *±*24 hours of *ICU* admission were reported.

Shock was defined as a cardiovascular *SOFA* score of 3 or more at *ICU* admission, equal to using a vasopressor (norepinephrine or epinephrine), combined with a lactate level of *>*2 mmol/L.

Community vs. hospital-acquired disease was distinguished based on hospital *LOS* before ICU admission. Community-acquired was defined as hospital *LOS ≤*2 days and hospital-acquired as *>*2 days.

Neutropenia was defined as a neutrophil count of *<*0.5×10^9^. Neutrophil count was analysed based on clinical indication and not routinely analysed in all patients. Leukopenia was defined as a White Blood Cell count (*WBC*)*<*3.5×10^9^.

Body temperature was obtained from the *SAPS −* 3 score and was the highest recorded within *±*1 hour of *ICU* admission.

See Supplement 2 for a complete list of variable definitions and data sources for Table 1.

### Biobank: blood sampling

Biobank blood samples were collected on *ICU* admission, typically at the time of routine blood sampling within one hour of arrival to the *ICU*. Samples were sent to the local hospital laboratory for centrifugation, aliquotation and freezing to −80°C before final storage. If freezing was delayed, samples were kept in a refrigerator until freezing. Blood samples were drawn, collected and stored for all patients without previous consent. A letter was sent to all patients who were alive 2-3 months after *ICU* care with information about the study. All participants could choose not to participate by contacting a study nurse or physician. By opting-out, all study data were deleted, and the blood samples were destroyed. This procedure was approved by the Ethical Committee at Lund University, Lund, Sweden (EPN 2015/256).

Patients were excluded if:

- samples were not drawn or were not matched to the correct patient
- patients were transferred between participating *ICU* s, and blood samples were not obtained anew after the transfer
- there was no consent

### Analysis of calprotectin and *CRP* from biobank samples

Frozen Ethylenediaminetetraacetic acid (*EDTA*)-plasma biobank samples were sent to the Department of Clinical Chemistry, Uppsala University Hospital, Uppsala, Sweden, where they were thawed and analysed. A Particle Enhanced Turbidimetric Assay (*PETIA*) methodology was used, with calprotectin reagents from Gentian AS (Moss, Norway) and CRP reagents from Abbott Laboratories (Abbott Park, IL, USA) on a Mindray BS380/BS430 chemistry analyser (Mindray, Shenzhen, China). Based on the sample handling times, additional samples were excluded due to calprotectin kinetics if one of the following criteria were met[19–21]:

- Stored at room temperature *>*72 hours after sampling
- Stored in refrigerator *>*7 days after sampling
- haemolysis in sample

### Statistical methods

Median values and Interquartile Range (*IQR*) were reported for continuous variables. For *SOFA*-scores, mean value and Standard Deviation (*SD*) were reported.

The Mann-Whitney U test was used to assess the difference between non-sepsis and sepsis in independent continuous variables. Differences in proportions were assessed using Pearson’s *χ*^2^ test. For all hypotheses testing, p-values *<*0.05 were considered significant.

Receiver Operating Characteristics (*ROC*) analysis with calculation of Area Under the Curve (*AUC*) was performed to assess the diagnostic capability of biomarkers, with sepsis as the outcome. Comparisons of *AUC*s was made using the DeLong test. For calculation of sensitivity, specificity, likelihood ratios and positive/negative predictive values, dichotomisation of calprotectin and *CRP* was based on optimal (Youden’s index) cutoffs derived from *ROC* curves, with high values considered positive tests.

Associations between calprotectin and other clinical variables were investigated using Spearman’s rank correlation coefficient (Spearman’s r). Locally Estimated Scatterplot Smoothing (*LOESS*) regression was used to visualise calprotectin as a function of CRP using the geom_smooth function of the ggplot2 package in R [22].

Patients with missing variables were excluded from analyses where the missing variable was used.

#### Subgroup analyses

Subgroup analyses were performed to explore differences in calprotectin levels between non-sepsis and sepsis and if the diagnostic performance of calprotectin differed in subgroups of *ICU* patients. The subgroups were chosen based on differences in disease course, illness severity and age to assess if calprotectin could be more useful in some subgroups. The subgroups were:

- community-acquired disease
- hospital-acquired disease
- neutropenic patients
- younger (*<*65 years) and older patients (*≥*65 years)
- patients with shock
- patients with *SOFA≥*2
- patients with suspected infection at *ICU* admission (blood cultures sampled and antibiotics administered)

Additional subgroup analyses were performed in microbiological subgroups of sepsis patients compared to non-sepsis patients. Calprotectin levels in bacteremic and non-bacteremic patients were also compared. For these subgroup analyses, differences in calprotectin levels were assessed with the Wilcoxon rank-sum test, with p*<*0.001 considered statistically significant, to compensate for multiple testing.

#### Sensitivity analyses

Sensitivity analyses were performed with altered criteria for organ failure or infection (as part of sepsis criteria) to investigate if the results were sensitive to which sepsis criteria were used:

- Organ failure defined as *SOFA≥*4
- Infection defined as only culture-positive with all culture-negative *ICU* patients defined as not infected
- Infection defined as obtained blood cultures and administered antibiotics (regardless of culture positivity or fulfilment of *LMCI*), according to the recommendation of the Sepsis-3-taskforce [1]

### Bias

A dropout analysis was performed to assess whether the biobank samples were missing at random.

The risk of confirmation bias when analysing the diagnostic capability of biomarkers made us change the infection criteria in this study, compared to the criteria we used in earlier studies of the same population [14, 16]. Biomarkers such as *CRP* and *WBC*, which were available to the clinician at *ICU* admission, are part of the clinical assessment leading to a decision to sample blood cultures and administer antibiotics, which was our single proxy criteria for infection in previous studies. This study has widened the infection criteria to include all culture-positive patients. For culture-negative sepsis, we applied previously used infection proxy criteria in combination with the assessment of infection according to the *LMCI*.

## Results

### Participants

Of 8360 *ICU* patients during the study period, 4732 were included. Sepsis criteria were fulfilled in 2071 (44%). The infection criteria included in the definition of sepsis were based on culture positivity in 81%, and the remaining 19% were culture negative with clinical criteria for infection. See Figure 1 for flow chart.

**Fig. 1:**
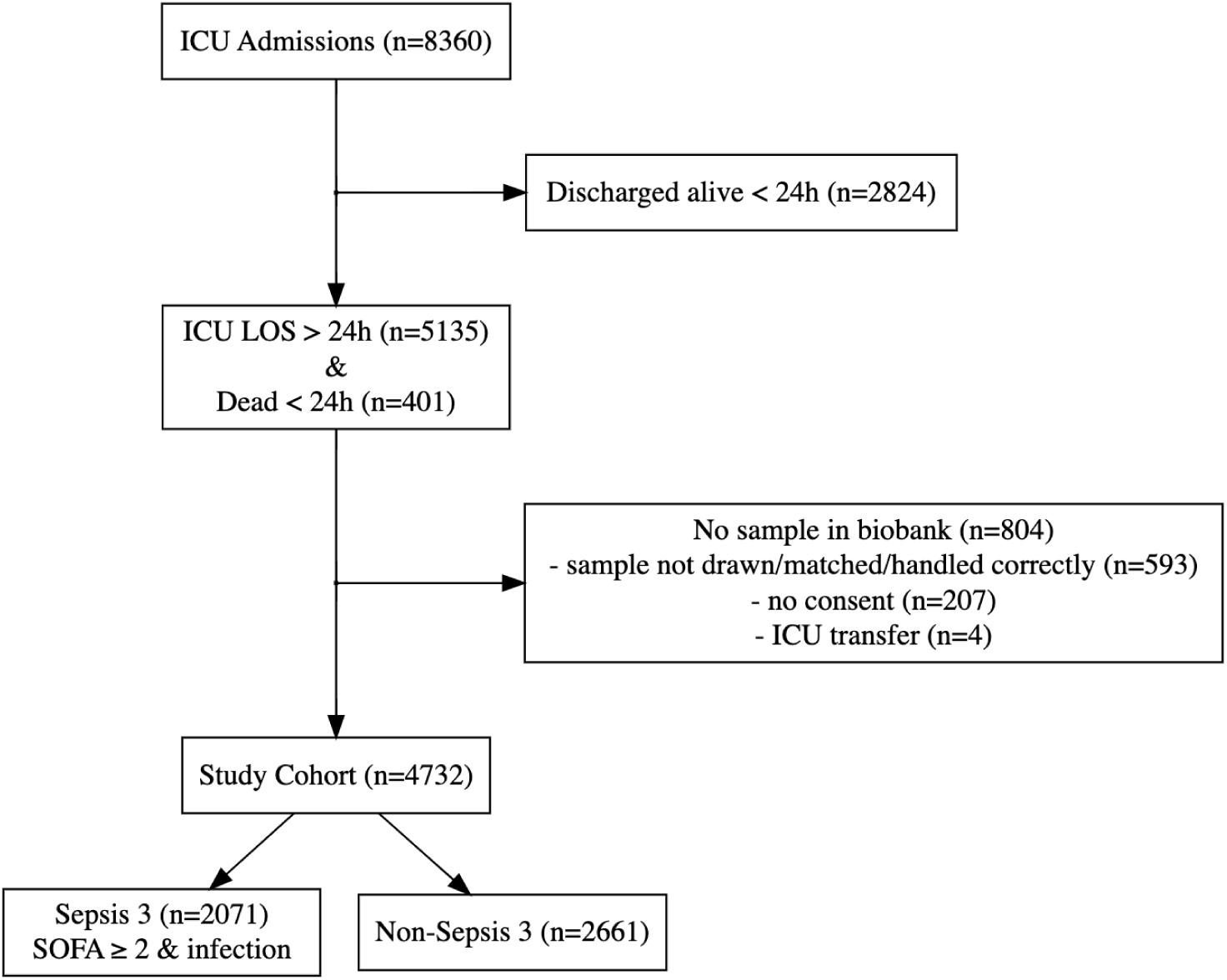
Flow chart of patient inclusion and exclusion. The included population had an *ICU LOS >*24 hours or died *≤*24 hours from *ICU* discharge, and had eligible biobank blood samples. Of those, 44% had sepsis. Infection criteria were culture positive *±* 48 hours or blood culture & antibiotics & probable infection according to LMCI. *ICU; intensive care unit. LOS; length of stay. LMCI; Linder-Mellhammar criteria of infection*.

### Baseline and outcome data

Table 1 describes the baseline and outcome data for the non-sepsis and sepsis cohorts. The sepsis cohort was slightly older (69 vs. 67 years, p*<*0.001), more severely ill (SOFA-score 8.3 vs 6.3, p*<*0.001) and had a higher proportion of shock (37% vs. 28%, p*<*0.001) than the non-sepsis cohort. There was no difference in the proportion of community/hospital-acquired disease or proportion being admitted directly from the Emergency Room (*ER*). Still, a smaller proportion of the sepsis cohort was admitted directly from the Operating Room (*OR*) than the non-sepsis cohort (8.4% vs. 20%, p*<*0.001). Regarding outcomes, the sepsis cohort had longer *ICU LOS* (3.2 vs. 2 days, p*<*0.001), higher 30-day mortality (33% vs. 28%, p*<*0.001) and more often needed Continous Renal Replacement Therapy (*CRRT*) (18% vs. 9.5%, p*<*0.001) and invasive mechanical ventilation (69% vs. 60%, p*<*0.001) during the *ICU* stay.

### Calprotectin and *CRP* levels in non-sepsis and sepsis

Calprotectin levels were higher in sepsis compared to non-sepsis (median 2.1 mg/L, IQR 1-4.3 and 1.3 mg/L, IQR 0.6-2.8, p*<*0.001). *CRP* was also elevated in sepsis compared to non-sepsis (median 95 mg/L, IQR 26-186 and 17 mg/L, IQR 3-69, p*<*0.001).

### *ROC* analysis and diagnostic testing of calprotectin and *CRP*

Calprotectin had a lower *AUC* (0.61, 95% CI 0.59-0.62) than *CRP* (0.72, 95% CI 0.71-0.74) in distinguishing sepsis from non-sepsis (p*<*0.001). Based on an optimal cutoff of 1.6 mg/L, the sensitivity was 59% and the specificity 57% for calprotectin to diagnose sepsis. For *CRP*, the optimal cutoff was 62 mg/L, resulting in a sensitivity of 62% and specificity of 73% (Table 2).

**Table 2:**
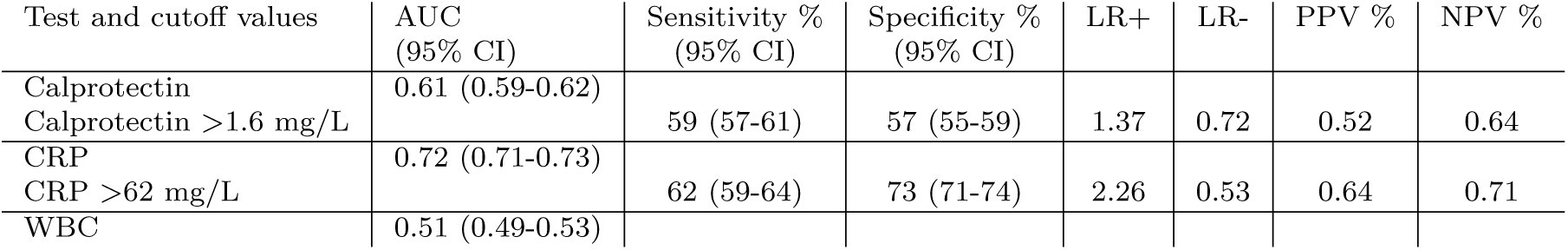
*AUC* of *ROC*-analysis and diagnostic testing of calprotectin, *CRP* and *WBC*. The outcome was confirmed sepsis, with non-sepsis as controls. Dichotomised calprotectin, *CRP* and *WBC* were based on an optimal cutoff (Youden’s index) from the *ROC* analysis, with high values considered positive tests. Diagnostic testing of *WBC* was not reported because of an *AUC* close to 0.5. *CRP; C-reactive protein. WBC; white blood cell count. AUC; area under the curve. CI; confidence interval. LR; likelihood ratio. PPV; positive predictive value. NPV; negative predictive value*.

### Association between calprotectin and *CRP*, *WBC* and *SOFA* score

There was a moderate correlation between calprotectin and *CRP* (Spearman’s r = 0.47) in the whole *ICU* population. Among the microbiological subgroups, the strongest association was in viral sepsis (Spearman’s r=0.57) and the weakest in gramnegative sepsis (Spearman’s r=0.40), see figure 3 and Table 2.5 in Supplement 1. The association was less pronounced with *WBC* count (Spearman’s r=0.28) and *SOFA* score (Spearman’s r=0.18).

### Subgroup analyses

The subgroup analyses showed higher median calprotectin levels in sepsis than non-sepsis across all subgroups, except in neutropenic patients, who also had markedly lower calprotectin levels (0.6 mg/L in non-sepsis and 1.0 mg/L in sepsis, p=0.91). The subgroup with a hospital-acquired disease had the highest calprotectin levels, both among non-sepsis (2.1 mg/L) and sepsis (2.6 mg/L) (Table 2.3 in Supplement 1). The ability of calprotectin to diagnose sepsis was lowest in patients with neutropenia (AUC 0.51) and hospital-acquired disease (AUC 0.55), and highest in patients with community-acquired disease and age*≥*65 years (AUC 0.62). The ability of calprotectin to diagnose sepsis remained inferior to *CRP* in all subgroups (Table 3).

**Table 3:** Subgroup analyses of the diagnostic capabilities of calprotectin and *CRP*. *ROC* analysis was performed with sepsis as the outcome and the *AUC* is displayed in the table. Comparison between *AUC* of calprotectin and *CRP* was evaluated with the deLong test. Community or hospital-acquired diseases were distinguished based on a hospital *LOS* under or over 48 hours before *ICU* admission. Shock was defined as vasopressor need and lactate *>*2 mmol/L. Suspected infection was defined as an obtained blood culture and administered antibiotics around the time of *ICU* admission. *ROC; Receiver Operating Curve. AUC; area under the curve. CRP; C-reactive protein. n; number. SOFA; sequential organ failure assessment*.

|  | n | AUC calprotectin | AUC CRP | p-value |
| --- | --- | --- | --- | --- |
| Included *ICU* population | 4732 | 0.61 | 0.72 | <0.001 |
| Community-acquired | 3536 | 0.62 | 0.75 | <0.001 |
| Hospital-acquired | 1196 | 0.55 | 0.63 | <0.001 |
| Neutropenic | 83 | 0.51 | 0.73 | 0.018 |
| Age<65 years | 1899 | 0.59 | 0.72 | <0.001 |
| Age≥65 years | 2833 | 0.62 | 0.72 | <0.001 |
| Shock | 1520 | 0.60 | 0.74 | <0.001 |
| SOFA≥2 | 4329 | 0.60 | 0.72 | <0.001 |
| Suspected infection | 2008 | 0.60 | 0.69 | <0.001 |

**Table 4:**
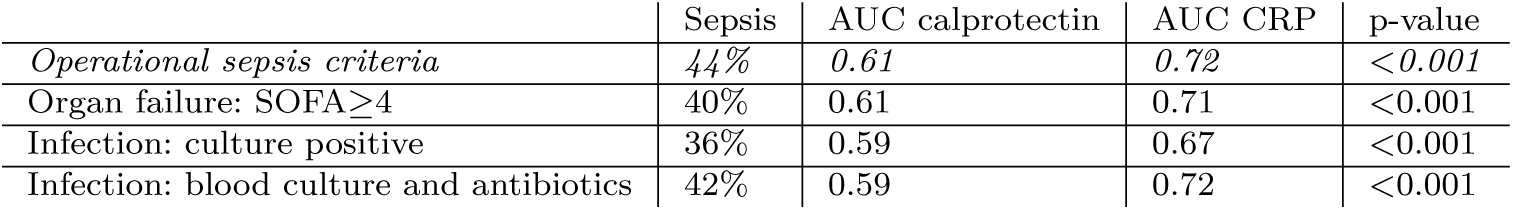
Sensitivity analyses with altered infection and organ failure criteria. The table displays the *AUC* of *ROC* analysis of each biomarker, with sepsis as outcome. The sensitivity analyses aimed to investigate the robustness of the main results when the criteria for organ failure and infection were altered as part of the sepsis criteria. The operational sepsis criteria of this study were: *SOFA* score of *≥*2 at *ICU* admission and infection. Infection was defined as positive culture or culture negativity but probable infection according to the *LMCI* infection criteria. The operational infection criteria were unchanged in the first analysis, but the SOFA criterion was raised to *≥*4. The infection criteria were altered in the last two sensitivity analyses, but the SOFA criterion of *≥*2 was unchanged. *ROC; Receiver Operating Curve. AUC; area under the curve. CRP; C-reactive protein. SOFA; sequential organ failure assessment*.

Among the microbiological subgroups, fungal sepsis had the highest calprotectin level (median 3.0 mg/L). Gram-positive and gram-negative sepsis generated the same level of calprotectin (median 1.9 mg/L), which was lower than for polymicrobial sepsis, and did not reach statistical significance for culture-negative sepsis (see figure 2 and Table 2.4 in Supplement 1).

**Fig. 2:**
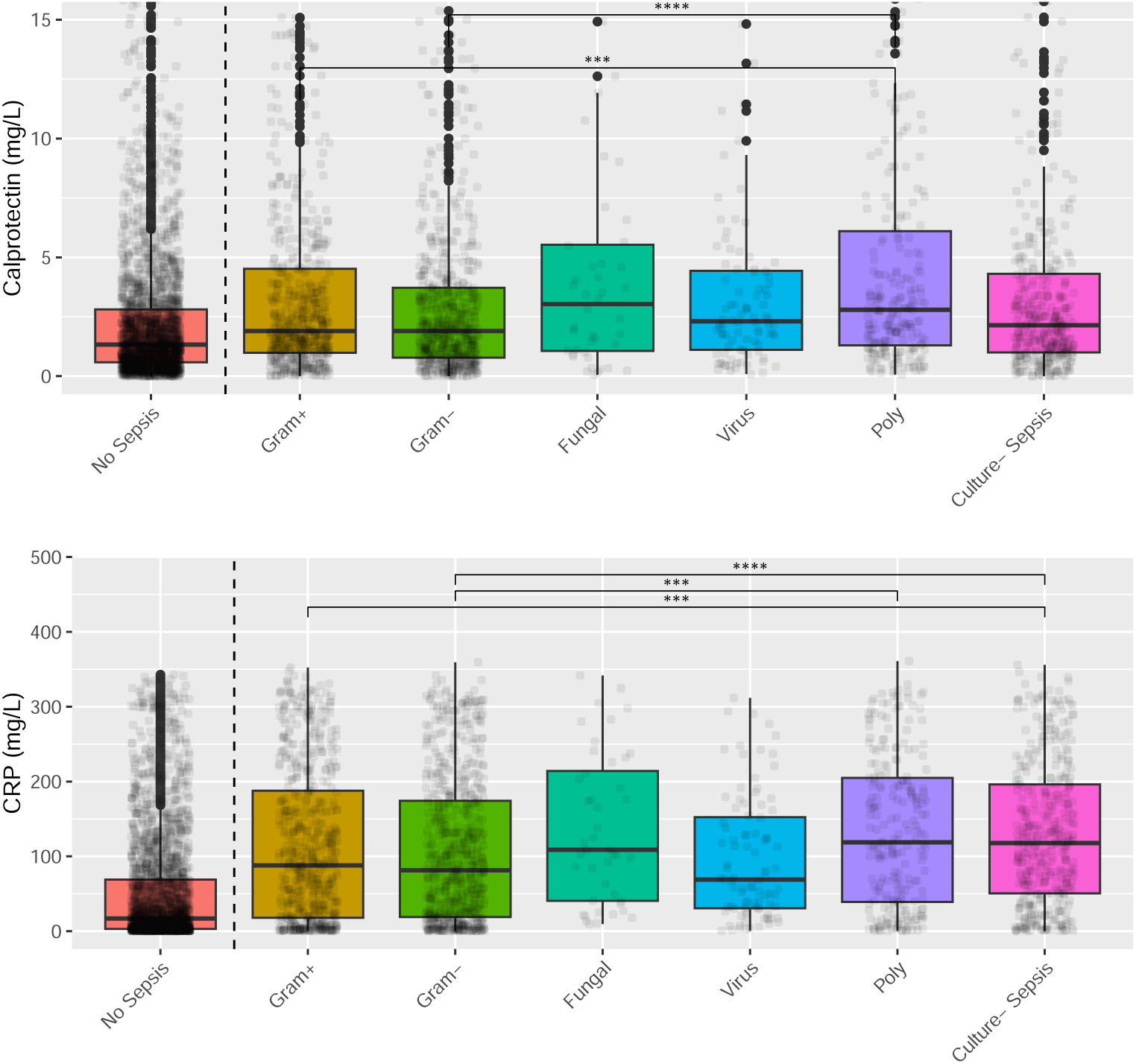
Boxplots of calprotectin and *CRP*. Sepsis patients were assigned to one of the microbiological subgroups based on relevant culture results within *±*48 hours of *ICU* admission. Only significances p *<*0.001 (***) and p *<*0.0001 (****) are shown to compensate for multiple testing. All microbiological subgrups of sepsis patients had higher calprotectin and *CRP* than non-sepsis, with a significance level *<*0.001. *CRP; C-reactive protein. +; positive. -; negative. Poly; polymicrobial*.

**Fig. 3:**
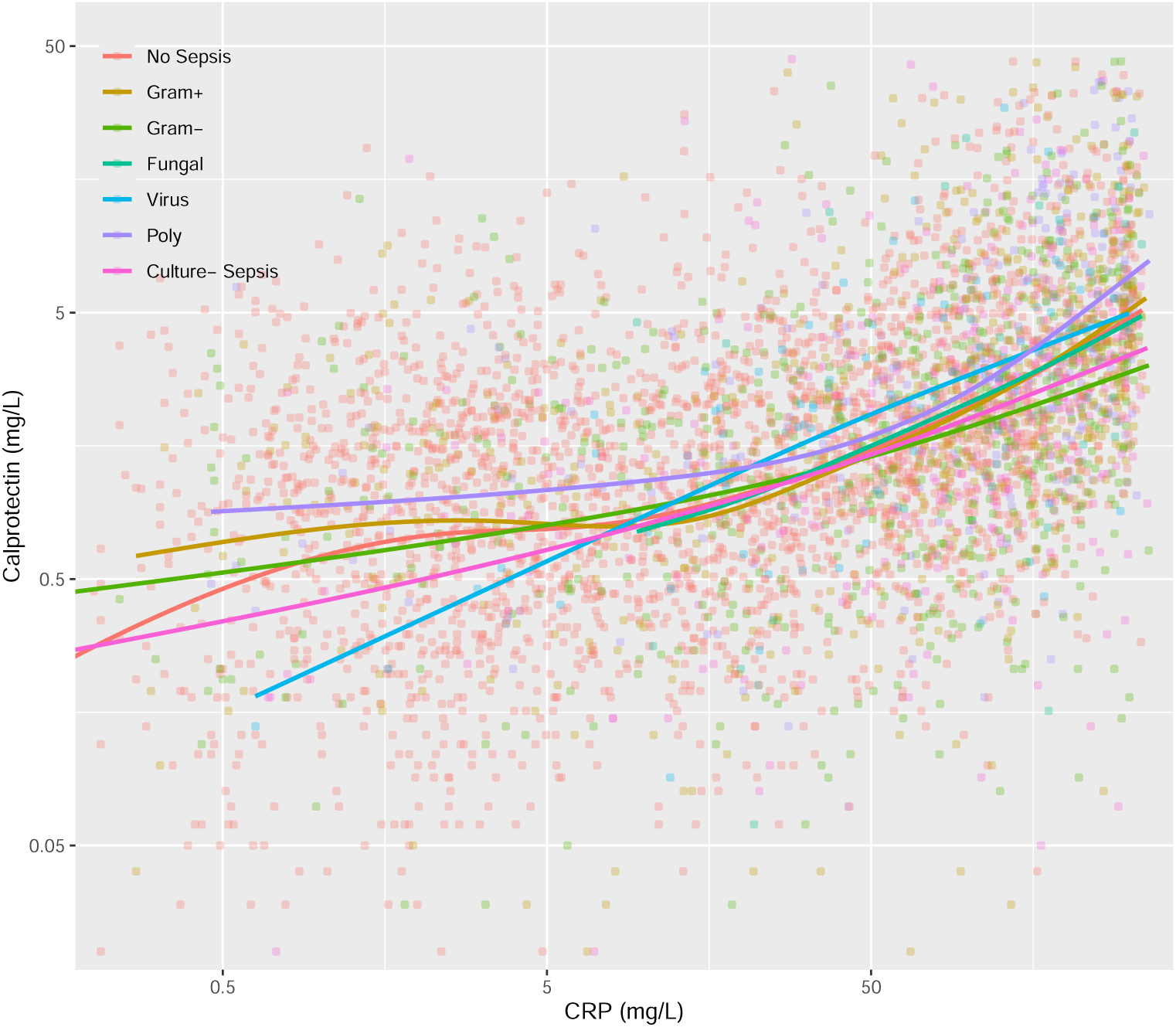
Calprotectin vs. *CRP*. The solid lines show the *LOESS* regression lines for the group with no sepsis and the microbiological subgroups of sepsis. The population’s global Spearman rank correlation of calprotectin and *CRP* was 0.47. *CRP; C-reactive protein. LOESS; locally estimated scatterplot smoothing*.

### Sensitivity analyses

The sensitivity analyses with altered criteria for organ failure and infection showed similar *AUC*s of calprotectin and *CRP* (*AUC* 0.59-0.61 for calprotectin and *AUC* 0.68-0.73 for *CRP*) to distinguish sepsis from non-sepsis, compared to the operational sepsis criteria used in the main analyses.

### Dropout analysis

Of the 5536 patients with *ICU LOS>*24h or who died within 24 hours, 804 (15%) did not have biobank blood samples and were not included in the study population (see flow chart in Figure 1). In comparison with the excluded patients, the study population had a higher degree of sepsis (44% vs. 32%, p*<*0.001) and shock (32% vs. 27%, p=0.004) and was more severely ill (*SOFA* score 7.2 vs. 6.4, p*<*0.001) but had lower 30-day mortality (30% vs 35%, p=0.008). A higher proportion of the included patients were admitted to the *ICU* directly from the *ER* (41% vs. 37%, p=0.012). Among the biomarkers routinely taken at *ICU* admission, lactate levels were higher (2.0 vs 1.8 mmol/L, p=0.031), but *CRP* levels were the same (43 vs 42 mg/L, p=0.28). See Table 2.1 in Supplement 1.

### Missing data

Due to the study design, no data for calprotectin or *CRP* were missing. See Supplement 2 for the proportion of missing values for the variables used in Table 1.

## Discussion

In this retrospective observational study of calprotectin and *CRP* as diagnostic markers of sepsis at ICU admission, the main finding was that calprotectin was inferior to *CRP* in diagnosing sepsis at *ICU* admission. Subgroup and sensitivity analyses followed a similar pattern.

Our study is the largest to date to examine calprotectin in the *ICU* setting, and the results do not support the use of calprotectin as a routine diagnostic marker of sepsis at *ICU* admission.

In our study, the diagnostic performance of calprotectin was poorer than in previous smaller studies[10, 11]. Larsson et al. found a slightly higher *AUC* of 0.67 for calprotectin to diagnose sepsis in a mixed *ICU* cohort but did not make a comparison with *CRP* [10]. However, the optimal cut-off value of 1.3 mg/L was similar to ours, using the same analysis method, *PETIA*. Differences in *AUC* may be explained by a smaller sample size (n=271) and different infection criteria to identify sepsis.

Jonsson et al. found an *AUC* of 0.76 for calprotectin to diagnose bacterial infection during the *ICU* stay, which was not significantly better than the *AUC* of 0.69 for *CRP* [11]. Noteworthy is that only patients without renal failure were included in the Jonsson study, and all patients with ongoing antibiotic treatment were excluded. In addition, most patients were admitted after multi-trauma with a high degree of Systemic Inflammatory Response Syndrome (*SIRS*), yielding a selected cohort of *ICU* patients. Additionally, a notably younger patient population was studied compared to our study (46 vs 68 years), and a bacterial infection alone was used as the outcome, not sepsis or other infections. These differences complicate a comparison to our study.

The proportion of patients with sepsis was higher in the present study compared to both comparable studies and in a general *ICU* population. This may be due to our inclusion criteria, with a lower proportion of sepsis among excluded patients discharged within the first 24 hours and a higher proportion among patients with biobank blood samples, as our dropout analysis revealed. Further, our wider infection criteria, which included positive microbiological cultures within four days, may have made a difference.

Among the microbiological subgroups, we did not find a difference in calprotectin levels between gram-positive and gram-negative bacteria, which is in discordance with the previous *ER* study, which found higher calprotectin levels in gram-positive sepsis[23]. This might be due to our patients being more severely ill and heterogeneous than an *ER* sepsis population with community-acquired sepsis. However, the disease severity in the present study, according to *SOFA*-score, had a very limited association with calprotectin levels. The only microbiological subgroup which had significantly higher calprotectin levels than the other groups was the polymicrobial subgroup, which could be due to a higher microbial load, a theory that prompts further investigation. In addition, bacteriemic sepsis patients had significantly higher calprotectin levels, probably due to a higher degree of neutrophil activation. Our finding that calprotectin levels are lowest in neutropenic patients further support previous studies suggesting that neutrophils are the most important source of systemic calprotectin[24, 25].

We also investigated the relationship between calprotectin and *CRP* levels. Only a moderate correlation between calprotectin and CRP was observed, suggesting that the two biomarkers reflect the activation of different immune and inflammatory pathways. While calprotectin is directly released by activated or dying neutrophils[26] and is mainly considered to be a marker of neutrophil function, CRP is produced in the liver due to IL-6 stimulation. As IL-6 has multiple cellular sources, including monocytes, macrophages, lymphocytes and several other cell types[27], IL-6 and CRP levels broadly reflect the systemic immune response. The different infection-related pathways of calprotectin and *CRP* are a potential explanation for the findings of our study.

A previous study indicated that *CRP* peaks later compared to calprotectin, and the more rapid kinetics of calprotectin might be more valuable in the early course of sepsis[23]. This difference in kinetics was not reflected in our subgroup analyses, however, since there was no difference in the ability of calprotectin to diagnose community vs. hospital-acquired sepsis.

Even though the diagnostic potential of calprotectin in sepsis was modest, a biomarker-guided therapy for subgroups of *ICU* patients with a high calprotectin at *ICU* admission could be of interest since calprotectin blockade has been shown to reduce the degree of inflammation and disease severity in animal models [28]. Also, our study cannot exclude the possibility that calprotectin dynamics, rather than admission calprotectin alone, might offer important prognostic information in these patients and guide treatment. In support of this hypothesis, we have recently shown that calprotectin measurement on day 7 post-admission offered much more robust information on disease course, complications and prognosis compared to admission calprotectin in severely ill Covid-19 patients[29].

### Strengths and limitations

Our study has several strengths. It is a multicentre study with a large patient population, reflecting the heterogeneity of *ICU* patients representative of an *ICU* population in a Scandinavian context. Therapeutic decisions were not influenced by calprotectin levels, allowing an unbiased evaluation of calprotectin as a sepsis biomarker. Also, we defined infection and sepsis through a rigorous method using results of microbiological testing, which is a strength since this information is often missing when assessing the likelihood of an infection at *ICU* admission. Compared to studies that use physician suspicion of infection alone as a proxy for infection, this is a strength.

The handling of biobank samples can lower or elevate measured levels of biomarkers. For calprotectin, neutrophils can be activated and secrete calprotectin in the sampling tube, leading to falsely high measured levels. For this reason, plasma is the preferred sample type for delayed analysis, as blood coagulation in the tube and subsequent platelet and neutrophil activation is avoided. We used EDTA-plasma and excluded samples with hemolysis or a delayed time to freezing to ensure that the measured calprotectin levels reflect the in vivo levels[19–21]. In addition, our chosen analysis method, *PETIA*, ensures that results are clinically applicable since *PETIA* is more suitable for clinical practice than other analysis methods, mainly because of shorter turnaround times and cost[30].

Our study also has limitations. The lack of generalisability to other *ICU* settings, particularly those with a different infection panorama and patient demographics, is a limitation.

Although the criteria for infection were chosen to minimise confirmation bias, there is still potential bias of the *CRP* analysis since the choice to obtain microbiological cultures and administer antibiotics is likely influenced by *CRP* levels. Therefore, our operational infection criteria could be influenced by *CRP* levels, which might partly explain the association between higher *CRP* and sepsis. However, this is a common limitation in studies of *CRP* as an infection marker. It is practically and ethically unfeasible to blind clinicians to *CRP* in the decision to investigate or treat for a presumed infection in current practice.

Another limitation is that we do not know where in the sepsis course calprotectin values were analysed. Our subgroup analysis of community vs. hospital-acquired disease did not affect calprotectin’s diagnostic capability, although calprotectin levels were higher in hospital-acquired disease. Patients could be very early or late in their sepsis course, which might affect calprotectin levels. This is a common limitation in sepsis studies since the time of sepsis onset is often difficult to determine.

Our study only assessed the value of calprotectin and *CRP* at admission, as serial samples were not available. As commented above, biomarker dynamics during the *ICU* stay might provide important information that could be used to guide patient care and assess prognosis. However, as the main purpose of our study was sepsis diagnosis and not prognosis, samples collected at admission are the most relevant.

## Conclusion

Calprotectin is inferior to *CRP* as a diagnostic marker of sepsis at *ICU* admission, and our findings do not support the use of calprotectin for this purpose.

## Supplementary information

Supplement 1 - method and results supplement. PDF file.
Supplement 2 - variable list, data sources and proportions missing for variables in Table 1. Excel file.

## Acronyms

AUC: Area Under the Curve
CRP: C-reactive Protein
CRRT: Continous Renal Replacement Therapy
EDTA: Ethylenediaminetetraacetic acid
EMR: Electronic Medical Records
ER: Emergency Room
ICU: Intensive Care Unit
IQR: Interquartile Range
LMCI: Linder-Mellhammar Criteria of Infection
LOESS: Locally Estimated Scatterplot Smoothing
LOS: Length of stay
OR: Operating Room
PETIA: Particle Enhanced Turbidimetric Assay
ROC: Receiver Operating Characteristics
SAPS – 3: Simplified Acute Physiology Score 3
SD: Standard Deviation
SIR: Swedish Intensive Care Registry
SIRS: Systemic Inflammatory Response Syndrome
SOFA: Sequential Organ Failure Assessment
STARD: Standards for Reporting Diagnostic accuracy studies
WBC: White Blood Cell count

## Acknowledgments

The authors would like to thank all collaborators in the SepCrit and Swecrit projects who have contributed to data collection.

## Declarations

### Ethics approval

The study protocol was approved by the Regional Ethical Review Board of Lund, Sweden (registration no. 2017/802 and 2015/267).

### Consent to participate

Following the ethics approval, consent for blood sampling to the biobank was presumed. After 2-3 months, survivors received a letter with information about the opportunity to opt out.

### Consent for publication

Not applicable.

### Availability of data and materials

The datasets generated and analysed during the current study are not publicly available due to limitations in the ethical approval of the study and data management policies of Region Skåne. Data might be available from the corresponding author at a reasonable request.

### Code availability

Available from the corresponding author on request.

### Authors’ contributions

ML, LM and AF designed the study. AF, AS, and AL funded the analyses. ML and SH collected data. ML and VSC cleaned and prepared the data. ML, SH and VSC performed statistical analyses. ML, SH and AF prepared the figures. ML wrote the draft manuscript. All authors read, provided critical revisions, and approved the final manuscript.

## References

[1] Singer M, Deutschman CS, Seymour CW, Shankar-Hari M, Annane D, Bauer M, et al. The third international consensus definitions for sepsis and septic shock (Sepsis-3). Jama. 2016;315(8):801–810.

[2] Vincent JL, Marshall JC, Ñamendys-Silva SA, François B, Martin-Loeches I, Lipman J, et al. Assessment of the worldwide burden of critical illness: the intensive care over nations (ICON) audit. The lancet Respiratory medicine. 2014;2(5):380–386.

[3] Póvoa P, Coelho L, Dal-Pizzol F, Ferrer R, Huttner A, Conway Morris A, et al. How to use biomarkers of infection or sepsis at the bedside: guide to clinicians. Intensive care medicine. 2023;49(2):142–153.

[4] Evans L, Rhodes A, Alhazzani W, Antonelli M, Coopersmith CM, French C, et al. Surviving sepsis campaign: international guidelines for management of sepsis and septic shock 2021. Intensive care medicine. 2021;47(11):1181–1247.

[5] Pierrakos C, Velissaris D, Bisdorff M, Marshall JC, Vincent JL. Biomarkers of sepsis: time for a reappraisal. Critical Care. 2020;24(1):1–15.

[6] Stríz I, Trebichavský I. Calprotectin—a pleiotropic molecule in acute and chronic inflammation. Physiol Res. 2004;53:245–253.

[7] Lipcsey M, Hanslin K, Stålberg J, Smekal D, Larsson A. The time course of calprotectin liberation from human neutrophil granulocytes after Escherichia coli and endotoxin challenge. Innate immunity. 2019;25(6):369–373.

[8] van Zoelen MA, Vogl T, Foell D, Van Veen SQ, van Till JW, Florquin S, et al. Expression and role of myeloid-related protein-14 in clinical and experimental sepsis. American journal of respiratory and critical care medicine. 2009;180(11):1098–1106.

[9] Bartáková E, Štefan M, Stráníková A, Pospíšilová L, Arientová S, Beran O, et al. Calprotectin and calgranulin C serum levels in bacterial sepsis. Diagnostic microbiology and infectious disease. 2019;93(3):219–226.

[10] Larsson A, Tydén J, Johansson J, Lipcsey M, Bergquist M, Kultima K, et al. Calprotectin is superior to procalcitonin as a sepsis marker and predictor of 30-day mortality in intensive care patients. Scandinavian Journal of Clinical and Laboratory Investigation. 2020;80(2):156–161.

[11] Jonsson N, Nilsen T, Gille-Johnson P, Bell M, Martling CR, Larsson A, et al. Calprotectin as an early biomarker of bacterial infections in critically ill patients: an exploratory cohort assessment. Critical Care and Resuscitation. 2017;19(3):205–213.

[12] Friberg H.: Swecrit Biobank - Blood Samples From Critically Ill Patients and Healthy Controls (SWECRIT). ClinicalTrials.gov.

[13] Cohen JF, Korevaar DA, Altman DG, Bruns DE, Gatsonis CA, Hooft L, et al. STARD 2015 guidelines for reporting diagnostic accuracy studies: explanation and elaboration. BMJ open. 2016;6(11):e012799.

[14] Lengquist M, Lundberg OHM, Spångfors M, Annborn M, Levin H, Friberg H, et al. Sepsis is underreported in Swedish intensive care units: A retrospective observational multicentre study. Acta Anaesthesiol Scand. 2020 09;64(8):1167–1176.

[15] Mellhammar L, Elén S, Ehrhard S, Bouma H, Ninck L, Muntjewerff E, et al. New, Useful Criteria for Assessing the Evidence of Infection in Sepsis Research. Critical care explorations. 2022;4(5):e0697.

[16] Lengquist M, Varadarajan A, Alestam S, Friberg H, Frigyesi A, Mellhammar L. Sepsis mimics among presumed sepsis patients at intensive care admission: a retrospective observational study. Publication in process. 2023;.

[17] Moreno RP, Metnitz PG, Almeida E, Jordan B, Bauer P, Campos RA, et al. SAPS 3—From evaluation of the patient to evaluation of the intensive care unit. Part 2: Development of a prognostic model for hospital mortality at ICU admission. Intensive care medicine. 2005;31:1345–1355.

[18] Vincent JL, Moreno R, Takala J, Willatts S, De Mendonça A, Bruining H, et al.: The SOFA (Sepsis-related Organ Failure Assessment) score to describe organ dysfunction/failure: On behalf of the Working Group on Sepsis-Related Problems of the European Society of Intensive Care Medicine (see contributors to the project in the appendix). Springer-Verlag.

[19] Pedersen L, Birkemose E, Gils C, Safi S, Nybo M. Sample type and storage conditions affect calprotectin measurements in blood. The Journal of Applied Laboratory Medicine. 2018;2(6):851–856.

[20] Mylemans M, Nevejan L, Van Den Bremt S, Stubbe M, Vander Cruyssen B, Moulakakis C, et al. Circulating calprotectin as biomarker in neutrophil-related inflammation: pre-analytical recommendations and reference values according to sample type. Clinica Chimica Acta. 2021;517:149–155.

[21] Blavnsfeldt ABG, Parkner T, Knudsen CS. Plasma calprotectin–preanalytical stability and interference from hemolysis. Scandinavian Journal of Clinical and Laboratory Investigation. 2022;82(5):349–355.

[22] Wickham H. ggplot2: Elegant Graphics for Data Analysis. Springer-Verlag New York; 2016. Available from: https://ggplot2.tidyverse.org.

[23] Christensen EE, Binde C, Leegaard M, Tonby K, Dyrhol-Riise AM, Kvale D, et al. Diagnostic accuracy and added value of infection biomarkers in patients with possible sepsis in the Emergency Department. Shock (Augusta, Ga). 2022;58(4):251.

[24] Åsberg A, Løfblad L, Felic A, Aune MW, Hov GG, Fagerli UM. Using blood calprotectin as a measure of blood neutrophils. Scandinavian Journal of Clinical and Laboratory Investigation. 2021;81(4):303–306.

[25] Cotoi OS, Dunér P, Ko N, Hedblad B, Nilsson J, Björkbacka H, et al. Plasma S100A8/A9 correlates with blood neutrophil counts, traditional risk factors, and cardiovascular disease in middle-aged healthy individuals. Arteriosclerosis, thrombosis, and vascular biology. 2014;34(1):202–210.

[26] Schiopu A, Cotoi OS. S100A8 and S100A9: DAMPs at the crossroads between innate immunity, traditional risk factors, and cardiovascular disease. Mediators of inflammation. 2013;2013(1):828354.

[27] Grebenciucova E, VanHaerents S. Interleukin 6: at the interface of human health and disease. Frontiers in immunology. 2023;14:1255533.

[28] Jakobsson G, Papareddy P, Andersson H, Mulholland M, Bhongir R, Ljungcrantz I, et al. Therapeutic S100A8/A9 blockade inhibits myocardial and systemic inflammation and mitigates sepsis-induced myocardial dysfunction. Critical Care. 2023;27(1):374.

[29] Didriksson I, Lengquist M, Spångfors M, Leffler M, Sievert T, Lilja G, et al. Increasing plasma calprotectin (S100A8/A9) is associated with 12-month mortality and unfavourable functional outcome in critically ill COVID-19 patients. Journal of Intensive Care. 2024;12(1):26.

[30] Nilsen T, Sunde K, Larsson A. A new turbidimetric immunoassay for serum calprotectin for fully automatized clinical analysers. Journal of Inflammation. 2015;12(1):1–8.

