## Supplement 1 for "Calprotectin as a sepsis diagnostic marker in critical care: a retrospective observational study"

### 1 Methods supplement

#### 1.1 Definition of clinically relevant pathogens in microbiological samples

Microbiological cultures were defined as clinically relevant if there was growth or detection of a pathogen from a culture taken within the stipulated time frame, regardless of the anatomical site of culturing. The following culture results were considered clinically irrelevant:

1. Yeast fungi from non-sterile anatomical sites (e.g. airways, lower urinary tract, skin lesions)
2. Potentially colonising bacteria of the upper or lower respiratory tract, if found in only one airway culture: Moraxella sp., coagulase-negative Staphylococci, viridans (alpha hemolytic) Streptococci
3. Potential skin contaminants found in only one blood culture: coagulase-negative Staphylococci, viridans (alpha hemolytic) Streptococci, micrococcus sp., Propionibacterium Acnes, Corynebacterium sp., Bacillus sp. [1]
4. Unspecific culture results (e.g. "gram-positive mixed flora", "vaginal flora", "skin flora", "anaerobic mixed flora") from non-sterile anatomical sites
5. Bacterial growth of Clostridium difficile, without detection of toxin[2]
6. Pneumococcus antigen tests from urinary samples

#### 1.2 Classification of pathogens

Pathogens were categorized into gram-positive bacteria, gram-negative bacteria, viruses, and fungi. Bacteria were further categorised by family or species. Atypical bacteria were considered gram-negative bacteria in this study. Viruses and fungi were categorized as seen below. *Sp; species.*

**Gram-negative bacteria**

- Acinetobacter sp.
- Bacteroides sp.
- Escherica coli
- Fusobacterium sp.
- Haemophilus influenzae
- Klebsiella sp.
- Moraxella sp.
- Neisseria meningitidis
- Other Enterobacter sp.
- Pseudomonas aeruginosa
- Stenotrophomonas maltophilia
- Other gram-negative bacteria
- Unspecified gram-negative bacteria
- Atypical bacteria

**Fungi**

- Candida sp.
- Pneumocystis sp.
- Other fungi

**Viruses**

- Influenza type A and B
- Other viruses

**Gram-positive bacteria**

- Bacillus sp.
- Clostridium difficile
- Coagulase-negative staphylococci
- Corynebacterium sp.
- Cutibacterium acnes
- Enterococcus faecalis
- Enterococcus faecium
- Lactobacillus sp.
- Mycobacterium sp.
- Staphylococcus aureus
- Streptococcus pneumoniae
- Streptococcus pyogenes
- Viridans streptococci
- Other Clostridium
- Other Enterococci
- Other Betastreptococci
- Other Streptococci
- Other gram-positive bacteria
- Unspecified gram-positive bacteria

#### 1.3 Stratification of patients with culture-positive sepsis into one of five subgroups

Microbiological cultures and samples were filtered by if they were taken within ±48 of ICU admission and if they were considered clinically relevant.

Based on the culture results, all sepsis patients were sorted into one of five categories. The five categories were gram-positive bacteria, gram-negative bacteria, fungal, viral and polymicrobial. The polymicrobial patient could have the following combinations of pathogens:

- Mixed bacteria (meaning cultures contain both gram-positive bacteria and gram-negative bacteria)
- Gram-negative bacteria + fungi or virus
- Gram-positive bacteria + fungi or virus
- Mixed bacteria + fungi or virus
- Fungi + virus

Most of the patients had cultures containing only one group of pathogens (i.e. gram-positive bacteria, gram-negative bacteria, fungi or viruses). However, about a third of the patients had cultures belonging to more than one of these groups. These were sorted as follows:

1. If the patient had a positive blood culture, the type of pathogen growing in the blood culture decided which group the patient belonged to. If more than one type of pathogen was found in blood cultures, the patient was allocated to the polymicrobial cohort. If the patient, in addition to positive blood cultures, tested positive for viruses (with any method), they were also categorised as polymicrobial. A few exceptions were made when there was a significantly higher occurrence of one type of pathogen type among the cultures from one patient.
2. If there were no blood cultures, the type of pathogen and the anatomic location it was sampled from were considered next. With the help of Infectious Diseases specialist LM (co-author), the following hierarchy was decided upon:
   (a) If primary pathogens (see below) were found in their respective anatomic location, the type of that pathogen decided which group the patient belonged to. If more than one type of primary pathogen was found, the patient was assigned to the polymicrobial group.
   (b) Other significant pathogens were considered if no primary pathogens were found.
   (c) Viruses were always considered relevant.
   (d) Fungi (other than candida sp. in upper respiratory specimen) were considered relevant when found alone in the lungs or the abdomen without other relevant pathogens.

**Primary pathogens:**

- Urinary tract: Escherica coli, Proteus sp, Klebsiella and other enterobacter sp.
- Respiratory tract: Streptococcus pneumoniae, Haemophilus influenzae, Legionella sp. Mycoplasma pneumoniae.
- All cultures from the central nervous system, bones, and joints
- Group A streptococcus and meningococcus sp: always significant regardless of anatomical site.

**Significant pathogens:**

- Enterococcus faecium in abdomen.
- Enterococcus faecalis in abdomen. When found in urine without other significant pathogens elsewhere.

- Pseudomonas sp. in airways or urine
- Staphylococcus Aureus in any other location besides the urinary tract unless it coexists with the primary pathogen in the respiratory tract.
- Group C and G streptococcus in airways, skin, and soft tissue.

**Non-significant pathogens (unless no other significant pathogens were found)**

- Enterococcus faecium in urine. Enterococcus faecium and faecalis in the respiratory tract.
- Moraxella, Coagulase negative staphylococcus, corynebacteria, acinetobacter, propionibacterium, stenotrophomonas sp.
- Staphylococcus Aureus in urine
- Group B streptococcus
- Viridans streptococcus

#### 1.4 All pathogens found in microbiological cultures/tests

S 1: All pathogens found in cultures and other microbiological tests sorted by gram stain (bacteria) and family or species (all pathogens). In some cases, the laboratory was unable to identify bacteria further than by general characteristics.

| Family or species | Pathogen |
|---|---|
| ***Gram-negative bacteria*** | |
| **Acinetobacter sp.** | Acinetobacter baumannii |
| | Acinetobacter baumannii-calcoaceticus complex |
| | Acinetobacter pittii |
| | Acinetobacter sp. |
| | Acinetobacter ursingii |
| **Bacteroides sp.** | Bacteroides caccae |
| | Bacteroides fragilis |
| | Bacteroides ovatus |
| | Bacteroides sp. |
| | Bacteroides thetaiota omicron |
| | Bacteroides uniformis |
| | Bacteroides vulgatus |
| **Escherichia Coli** | Escherichia coli |
| **Fusobacterium sp.** | Fusobacterium gonidia formans |
| | Fusobacterium necrophorum |
| | Fusobacterium nucleatum |
| | Fusobacterium sp. |
| **Haemophilus** | Haemophilus influenzae |
| | Haemophilus parainfluenzae |
| | Haemophilus sp. |
| **Klebsiella sp.** | Klebsiella-Enterobacter sp. |
| | Klebsiella aerogenes |
| | Klebsiella oxytoca |

| | |
|---|---|
| | Klebsiella oxytoca group |
| | Klebsiella pneumoniae |
| | Klebsiella sp. |
| **Moraxella sp.** | Moraxella catarrhalis |
| | Moraxellanon liquefaciens |
| **Neisseria meningitidis** | Neisseria meningitidis |
| **Other enterobacter** | Citrobacter freundii |
| | Citrobacter koseri |
| | Citrobacter sp. |
| | Enterobacter asburiae |
| | Enterobacter cloacae |
| | Enterobacter cloacae complex |
| | Enterobacter sp. |
| | Enterobacterales |
| | Hafnia alvei |
| | Morganella morganii |
| | Pantoea agglomerans |
| | Pantoea sp. |
| | Proteus mirabilis |
| | Proteus penneri |
| | Proteus sp. |
| | Proteus vulgaris |
| | Proteus vulgaris group |
| | Providencia rettgeri |
| | Salmonella dublin |
| | Salmonella enteritidis |
| | Salmonella florida |
| | Salmonella group O10 |
| | Salmonella sp. |
| | Salmonella subsp. |
| | Salmonella weltevreden |
| | Serratia liquefaciens |
| | Serratia marcescens |
| | Serratia rubidaea |
| | Serratia sp. |
| **Pseudomonas aeruginosa** | Pseudomonas aeruginosa |
| **Stenotrophomonas maltophilia** | Stenotrophomonas maltophilia |
| **Other gram-negative bacteria** | Achromobacter sp. |
| | Achromobacter xylosoxidans |
| | Aeromonas hydrophila |
| | Aeromonas sp. |
| | Alcaligenes faecalis |
| | Alloscardovia omnicolens |
| | Bordetella bronchiseptica |
| | Burkholderia cenocepacia |
| | Burkholderia cepacia |
| | Burkholderia cepacia complex |
| | Burkholderia gladioli |

| | |
|---|---|
| | Campylobacter jejuni/coli |
| | Campylobacter ureolyticus |
| | Capnocytophaga sp. |
| | Chryseobacterium gleum |
| | Chryseobacterium indologenes |
| | Chryseobacterium sp. |
| | Comamonas testosteroni |
| | Cronobacter sakazakii |
| | Cyberlindner afabianii |
| | Delftia acidovorans |
| | Dermabacter hominis |
| | Dialister pneumosintes |
| | Eikenella corrodens |
| | Elizabethkingia meningoseptica |
| | Elizabethkingia miricola |
| | Elizabethkingia sp. |
| | Hungatella hathewayi |
| | Leclercia adecarboxylata |
| | Lichtheimia corymbifera |
| | Neisseria bacilliformis |
| | Neisseria sp. |
| | Neisseria subflava |
| | Ochrobactrum anthropi |
| | Ochrobactrum sp. |
| | Pasteurella multocida |
| | Porphyromonas endodontalis |
| | Porphyromonas somerae |
| | Porphyromonas sp. |
| | Prevotella bivia |
| | Prevotella denticola |
| | Prevotella melaninogenica |
| | Prevotella nigrescens |
| | Prevotella oralis |
| | Prevotella oris |
| | Prevotella sp. |
| | Pseudomonas sp. |
| | Sphingomonas paucimobilis |
| | Sphingomonas sp. |
| | Sutterella wadsworthensis |
| | Veillonella parvula |
| | Veillonella sp. |
| | Vibrio sp. |
| | Vibrio vulnificus |
| **Atypical bacteria** | Legionella longbeachae |
| | Legionella pneumophila |
| | Legionella sp. |
| | Mycoplasma hominis |
| | Chlamydophila psittaci (DNA) |

| | |
|---|---|
| **Unspecified gram-negative bacteria** | Anaerobe gram-negative rods |
| | Gram-negative mixed flora |
| | Gram-negative mixed flora, e.g proteus |
| | Gram-negative mixed flora, e.g pseudomonas |
| | Gram-negative mixed environmental bacteria |
| ***Gram-positive bacteria*** | |
| **Bacillus sp.** | Bacillus cereus |
| | Bacillus cereus complex |
| | Bacillus sp. |
| **Clostridium Difficile** | Clostridium difficile toxin Vidas |
| | Clostridium difficile bacteria |
| **Coagulase-negative staphylococci** | Staphylococcus capitis |
| | Staphylococcus epidermidis |
| | Staphylococcus haemolyticus |
| | Staphylococcus hominis |
| | Staphylococcus lugdunensis |
| | Staphylococcus pasteuri |
| | Staphylococcus pettenkoferi |
| | Staphylococcus pseudintermedius |
| | Staphylococcus saprophyticus |
| | Staphylococcus schleiferi |
| | Staphylococcus simulans |
| | Staphylococcus sp. (coagulase-negative) |
| **Corynebacterium sp.** | Corynebacterium pseudodiphtheriticum |
| | Corynebacterium sp. |
| | Corynebacterium striatum |
| | Corynebacterium tuberculostearicum |
| **Cutibacterium acnes** | Cutibacterium acnes |
| **Enterococcus faecalis** | Enterococcus faecalis |
| **Enterococcus faecium** | Enterococcus faecium |
| **Lactobacillus sp.** | Lactobacillus gasseri |
| | Lactobacillus plantarum |
| | Lactobacillus rhamnosus |
| | Lactobacillus sp. |
| | Lactococcus sp. |
| **Mycobacteria** | Mycobacteria seen in microscope |
| | Mycobacterium tuberculosis |
| **Staphylococcus aureus** | Staphylococcus aureus |
| **Streptococcus pneumoniae** | Streptococcus pneumoniae |
| | Likely pneumococci |
| **Streptococcus pyogenes** | Streptococcus pyogenes(GAS) |
| **Viridans streptococci** | Streptococcus anginosus |
| | Streptococcus anginosus complex |
| | Streptococcus massiliensis |
| | Streptococcus mitis |
| | Streptococcus mitis complex |
| | Streptococcus mutans complex |
| | Streptococcus oralis |

| | |
|---|---|
| | Streptococcus parasanguinis |
| | Streptococcus salivarius complex |
| | Streptococcus sanguinis complex |
| | Streptococcus vestibularis |
| **Other Clostridium** | Clostridium aldenense |
| | Clostridium butyricum |
| | Clostridium cadaveris |
| | Clostridium innocuum |
| | Clostridium paraputrificum |
| | Clostridium perfringens |
| | Clostridium ramosum |
| | Clostridium septicum |
| | Clostridium sp. |
| | Clostridium symbiosum |
| | Clostridium tertium |
| **Other enterococci** | Enterococcus avium |
| | Enterococcus casseliflavus |
| | Enterococcus durans |
| | Enterococcus gallinarum |
| | Enterococcus gallinarum group |
| | Enterococcus hirae |
| | Enterococcus raffinosus |
| | Enterococcus sp. |
| **Other beta streptococci** | Beta streptococci group C |
| | Beta streptococci group G |
| | Streptococcus agalactiae (group B) |
| | Streptococcus dysgalactiae |
| | Streptococcus dysgalactiae (group C) |
| | Streptococcus dysgalactiae (group G) |
| **Other streptococci** | Streptococcus bovis |
| | Streptococcus bovis complex |
| | Streptococcus sp. |
| **Other gram-positive bacteria** | Actinomyces odontolyticus |
| | Actinomyces sp. |
| | Actinomyces turicensis |
| | Actinotignum sanguinis |
| | Actinotignum schaalii |
| | Actinotignum sp. |
| | Aerococcus sanguinicola |
| | Aerococcus sp. |
| | Aerococcus urinae |
| | Aggregatibactera phrophilus |
| | Aggregatibacter sp. |
| | Alistipesonder donki |
| | Anaerococcus sp. |
| | Aureobasidium pullulans |
| | Bifidobacterium breve |
| | Bifidobacterium sp. |

| | |
|---|---|
| | Brevundimonas aurantiaca |
| | Brevundimonas sp. |
| | Cutibacterium avidum |
| | Cutibacterium granulosum |
| | Cutibacterium sp. |
| | Desulfovibrio desulfuricans |
| | Dialister sp. |
| | Difteroida stavar |
| | Eggerthella lenta |
| | Eggerthella sp. |
| | Eggerthiaca tenaformis |
| | Enterocloster clostridioformis |
| | Eubacterium sp. |
| | Facklamia hominis |
| | Finegoldi amagna |
| | Galactomyces candidus |
| | Gemella haemolysans |
| | Gemella morbillorum |
| | Gemella sp. |
| | Globicatella sp. |
| | Grampositiva kocker |
| | Granulicatella adiacens |
| | Granulicatella sp. |
| | Listeria innocua |
| | Listeria monocytogenes |
| | Massilia sp. |
| | Micrococcus luteus |
| | Micrococcus sp. |
| | Nocardia sp. |
| | Odoribacter splanchnicus |
| | Paenibacillus sp. |
| | Paeniclostridium sordellii |
| | Parabacteroides distasonis |
| | Parabacteroides sp. |
| | Paracoccus sp. |
| | Parvimonas micra |
| | Peptoniphilus harei |
| | Peptoniphilus sp. |
| | Peptostreptococcus anaerobius |
| | Peptostreptococcuss tomatis |
| | Pichia cactophila |
| | Pichia norvegensis |
| | Rothia mucilaginosa |
| | Rothia sp. |
| | Ruminococcus gnavus |
| | Solobacterium moorei |
| | Staphylococcus warneri |
| | Weissella sp. |

| | |
|---|---|
| **Unspecified gram-positive bacteria** | Anaerobe gram-positive rods |
| | Gram-positive cocci, likely streptococci or enterococci |
| | Gram-positive rods |
| | Gram-positive mixed flora |
| ***Unspecified gram stain*** | |
| **Unspecified** | Anaerobe bacteria |
| | Anaerobe mixed flora |
| | Bacteria |
| | Mixed flora |
| | Mixed flora, e.g proteus |
| | Mixed flora, e.g pseudomonas |
| | Fecal flora |
| | Skin flora |
| | Oral flora |
| | Oropharyngeal flora |
| | Vaginal flora |
| ***Fungi*** | |
| **Candida** | Candida albicans |
| | Candida dubliniensis |
| | Candida glabrata |
| | Candida inconspicua |
| | Candida krusei |
| | Candida parapsilosis |
| | Candida sp. |
| | Candida tropicalis |
| | Candida utilitis |
| **Pneumocystis** | Pneumocystis |
| **Other fungi** | Aspergillus fumigatus |
| | Aspergillus nidulans |
| | Aspergillus sp. |
| | Aspergillus terreus |
| | Clavispora lusitaniae |
| | Diutinaca tenulata |
| | Diutinaca tenulata |
| | Fusarium solani |
| | Fusarium sp. |
| | Geotrichum sp. |
| | Yeast fungi |
| | Yeast fungi, not C. albicans |
| | Kluyveromyces marxianus |
| | Magnusiomyces capitatus |
| | Malassezia sp. |
| | Meyerozyma guilliermondii |
| | Mold fungi |
| | Mucor circinelloides |
| | Rhizopus microsporus |
| | Saccharomyces cerevisiae |
| | Thread fungus |

| | |
|---|---|
| | Trichosporona sahii |
| | Wickerhamomyces anomalus |
| ***Viruses*** | |
| **Influenza** | Influenzae A |
| | Influenzae B |
| **Other viruses** | Varicella zoster virus |
| | RS-virus type B |
| | RS-virus type A |
| | Rhinovirus |
| | Parainfluenzae |
| | Norovirus I |
| | Norovirus II |
| | Metapneumovirus |
| | Herpes simplex virus 1 |
| | Enterovirus |
| | Coronavirus |
| | Cytomegalovirus |
| | Adenovirus |

### 2 Results supplement

#### 2.1 Dropout analysis

Table of dropout analysis of included and excluded patients. The excluded patients did not have blood samples in the biobank. CRP in this table is from routine laboratory analysis. All variables, except outcomes, are at ICU admission. CRRT and mechanical ventilarion are during the ICU stay. *ICU; intensive care unit. LOS; length of stay. ER; emergency room. IQR; interquartile range. SD; standard deviation. CRP; C-reactive protein. WBC; white blood cell count. SAPS-3; simplified acute physiology score-3. SOFA; sequential organ failure assessment. CRRT; continuous renal replacement therapy.*

| | Included | Excluded | p-value |
|---|---|---|---|
| n | 4732 | 804 | |
| Age, years (median [IQR]) | 68 [56-75] | 66 [50-74] | <0.001 |
| Male sex (%) | 2856 (60) | 467 (58) | 0.24 |
| ICU admission >48h from hospital admission (%) | 1196 (25) | 264 (33) | <0.001 |
| Admitted from ER (%) | 1956 (41) | 294 (37) | 0.012 |
| Biomarkers | | | |
| C-reactive protein, mg/L (median [IQR]) | 43 [7-144] | 42 [7-128] | 0.28 |
| Lactate, mmol/L (median [IQR]) | 2.0 [1.2-4.1] | 1.8 [1-4.4] | 0.031 |
| WBC, $10^9$/L (median [IQR]) | 12.9 [8.7-18] | 12.6 [9.1-18.7] | 0.4 |
| Physiological parameters | | | |
| Sepsis (%) | 2071 (44) | 260 (32) | <0.001 |
| Shock (%) | 1520 (32) | 207 (27) | 0.004 |
| SAPS-3 and SOFA score at ICU admission | | | |
| SAPS-3 score (median [IQR]) | 64 [52-75] | 62 [49-74] | 0.008 |
| SOFA score (mean (SD)) | 7.2 (4) | 6.4 (4.2) | <0.001 |
| Outcomes | | | |
| ICU LOS (median [IQR]) | 2.5 [1.5-4.8] | 2.2 [1.2-4.4] | <0.001 |
| ICU mortality (%) | 871 (18.4) | 175 (21.8) | 0.028 |
| 30-day mortality (%) | 1435 (30) | 252 (35) | 0.008 |
| CRRT (%) | 627 (13) | 87 (11) | 0.065 |
| Invasive mechanical ventilation (%) | 3029 (64) | 479 (60) | 0.018 |

#### 2.2 Sample handling times

The median time from ICU admission to freezing of the biobank sample was 1.6 hours (IQR 1.3-2.1 hours). Among the samples with missing data at the time of freezing (n=45), the median time from ICU admission to the refrigerator was 2.2 hours (IQR 1.6-3.0).

#### 2.3 Calprotectin levels in subgroups of ICU patients.

Calprotectin levels are displayed as median (IQR) for the non-sepsis and sepsis groups within the subgroups of ICU patients. Community or hospital-acquired disease was defined as a hospital LOS under respective over 48 hours before ICU admission. Shock was defined as vasopressor need and a lactate >2 mmol/L. Suspected infection was defined as an obtained blood culture and administered antibiotics around the time of ICU admission. *ICU; intensive care unit. LOS; length of stay. n; number. IQR; interquartile range.*

| | n | Sepsis (%) | Non-sepsis median (IQR) | Sepsis median (IQR) | p-value |
|---|---|---|---|---|---|
| *Full ICU cohort* | *4732* | *44%* | *1.3 (0.6-2.8)* | *2.1 (1.0-4.3)* | <0.001 |
| Community-acquired | 3536 | 43% | 1.1 (0.5-2.4) | 1.9 (0.9-3.9) | <0.001 |
| Hospital-acquired | 1196 | 45% | 2.1 (1.0-4.5) | 2.6 (1.2-5.6) | 0.003 |
| Neutropenic | 83 | 82% | 0.6 (0.3-3) | 1.0 (0.2-3.5) | 0.91 |
| Age<65 years | 1899 | 46% | 1.3 (0.6-2.6) | 2.1 (1-4.1) | <0.001 |
| Age≥65 years | 2833 | 41% | 1.4 (0.5-3.1) | 2 (0.9-4.6) | <0.001 |
| Shock | 1520 | 51% | 1.6 (0.8-3.1) | 2.3 (1.1-4.4) | <0.001 |
| SOFA≥2 | 4329 | 49% | 1.4 (0.6-2.9) | 2.1 (1-4.3) | <0.001 |
| Suspected infection | 2008 | 77% | 1.4 (0.6-3.2) | 2.2 (1-4.7) | <0.001 |

#### 2.4 Calprotectin levels in microbiological subgroups of sepsis patients.

The microbiological subgroups of sepsis patients were based on relevant culture results in ±48 hours from ICU admission. Patients were assigned to only one of the six microbiological sepsis subgroups (gram-positive, gram-negative, fungal, virus, polymicrobial and culture-negative). All microbiological sepsis subgroups had higher calprotectin levels than the non-sepsis group ($p<0.05$). When the sepsis group was divided into bacteriemia and non-bacteriemia, the difference in calprotectin levels was significant with a p-value<0.001. *n; number. IQR; interquartile range.*

| Microbiological subgroup | n | Calprotectin, median (IQR) |
|---|---|---|
| *Non-sepsis* | *2661* | *1.3 (0.6-2.8)* |
| Gram-positive | 613 | 1.9 (1.0-4.5) |
| Gram-negative | 697 | 1.9 (0.8-3.7) |
| Fungal | 44 | 3.0 (1-5.6) |
| Virus | 92 | 2.3 (1.1-4.5) |
| Polymicrobial | 236 | 2.8 (1.3-6.2) |
| Culture-negative | 389 | 2.1 (1-4.3) |
| Bacteriemic | 539 | 2.6 (1.2-5.7) |
| Non-bacteriemic | 1532 | 1.9 (0.9-3.8) |

#### 2.5 Association between calprotectin and CRP in microbiological subgroups of sepsis patients.

The Spearman's r is displayed for the non-sepsis patients and for each microbiological sugroup of sepsis patients. All correlations were statistically significant with p<0.05. The number (n) of patients in each subgroup is the same as in table 2.4 above. *CRP; C-reactive protein.*

| Microbiological subgroup | Spearman's r |
|---|---|
| *Full ICU cohort* | 0.47 |
| *Non-sepsis* | 0.41 |
| Gram-positive | 0.51 |
| Gram-negative | 0.40 |
| Fungal | 0.41 |
| Virus | 0.57 |
| Polymicrobial | 0.54 |
| Culture-negative | 0.41 |

### References

[1] Alcorn K, Meier F, Schifman R. Blood Culture Contamination: Q-tracks 2013. College of American Pathologists. 2013.

[2] Crobach M, Planche T, Eckert C, Barbut F, Terveer E, Dekkers O, et al. European Society of Clinical Microbiology and Infectious Diseases: update of the diagnostic guidance document for Clostridium difficile infection. Clinical microbiology and infection. 2016;22:S63-81.
